# Application of the Concept ‘Avoidable Mortality’ in Assessing the Socioeconomic Status related Inequalities in Health: A Scoping Review

**DOI:** 10.1101/2023.09.07.23295200

**Authors:** Anousheh Marouzi, Charles Plante, Cory Neudorf

## Abstract

**Introduction:** Avoidable mortality is widely used by public health researchers to measure population health, and many related methodologies have been proposed for doing so. This scoping review presents a comprehensive view of global peer-reviewed and grey literature exploring the association between socioeconomic status (SES) and avoidable mortality.

**Methods:** We searched Ovid Medline, Scopus, and Web of Science to find articles that investigated SES inequalities in avoidable mortality. We limited our review to articles in English published between 2000 and 2020. For grey literature, we searched leading global and Canadian health information websites. We extracted data on different study characteristics, avoidable mortality definition, SES indicator, method of analysis of the association between avoidable mortality and SES, and main findings of the studies.

**Results:** We identified 34 articles to review, including 29 scientific papers and 5 grey literature documents. The findings of the selected articles consistently indicate a negative association between SES and avoidable mortality rates. Studies have not all used the same definitions of avoidable mortality or SES nor operationalized them in the same way.

**Conclusion:** Our review highlights the absence of a globally standard definition in avoidable mortality health equity research. Additional work to establish a standardized definition is crucial for supporting global comparability.

## Introduction

Addressing health inequities requires improved measurement, monitoring, and reporting of population health status and its association with social determinants of health.^1–3^ Avoidable mortality, a subset of premature mortality, is widely used by public health researchers to measure population health, and many related methodologies have been proposed for doing so. This scoping review presents a comprehensive view of global peer-reviewed and grey literature exploring the association between socioeconomic status (SES) and avoidable mortality. The studies tend to vary not only in how they define avoidable mortality but also in how they measure SES and the association between them. We also report on how these studies do or do not standardize their results for age and sex and encapsulate their main findings with respect to SES inequalities. Our aim is to facilitate a more widespread and in-depth analysis of inequities in avoidable mortality by consolidating these resources in one location.

Avoidable mortality is a relatively new concept, with corresponding methodologies still under refinement. In 1976, the Working Group on Preventable and Manageable Diseases, chaired by David D. Rutstein, introduced the new concept of “unnecessary untimely death” as a health outcome indicator designed to evaluate the quality of medical care.^4^ Subsequently, an American research group empirically applied this concept to “demonstrate the usefulness of this approach” as a measure of healthcare quality.^5,6^ The term “avoidable mortality” came into use following a study by Charlton et al., investigating geographical variation in mortality across England and Wales.^7^ While premature death is defined as mortality that happens before a certain age, for example before 75 years of age,^8^ “avoidable mortality” refers to premature deaths that could have been avoided in the presence of timely and effective health and social policies and public health interventions aimed at addressing the social determinants of health or reducing risk factors contributing to ill health.^8^ Nevertheless, researchers do not adopt a uniform approach to conceptualizing “avoidable mortality,” and it is not always clear whether the terms “avoidable,” “amenable,” “preventable,” and “unnecessary” causes of death are being used interchangeably in the literature.^9,10^ Over time, researchers have refined and updated the list of avoidable causes of death used to classify mortality events as avoidable to suit the situational context of their research.^8,11–16^

Avoidable mortality is an important summary measure of overall population health status,^8^ and one of its many applications has been as an indicator of health equity.^5,17^ Researchers have used avoidable mortality to investigate health inequity with the goal of informing political decisions trying to address health inequity.^11,18–22^ Socioeconomic status (SES) is one of the most important “fundamental causes” of disparities in morbidity and mortality.^23,24^ People with lower SES have access to a limited range of resources, such as money, knowledge, prestige, social capital, and power, which could affect their health through many pathways; lack of these resources exposes individuals to a higher risk of mortality.^23^ These deaths could be avoided by appropriate means such as timely and effective treatment or the application of health and social policies.

## Methods

We used Arksey and O’Malley’s framework^25^ to conduct this scoping review in five stages:

### 1. Identification of studies

Our central question in this review was “How have avoidable mortality and inequalities therein been defined and operationalized in studies investigating SES inequalities in avoidable mortality?” To answer this question, we developed a search strategy (see Appendix A) to find relevant articles by searching Ovid Medline, Scopus, and Web of Science, narrowing down the results to English language and publication dates from 2000 to 2020. We omitted PubMed in our search since the Ovid Medline interface allows a more focused search, excluding citations such as “in process” and “ahead of print” articles,^26^ which were not of interest to the current review. All the searches were conducted on June 18th, 2020. For grey literature, since our research team is located in Canada, we were especially interested in Canadian literature. We conducted searches on leading Canadian health information websites and several leading international websites. We searched Google, Google Scholar, The World Health Organization (WHO),^27^ United Nations,^28^ Organization for Economic Co-operation and Development (OECD),^29^ and Public Health Agency of Canada (PHAC)^30^ restricting our review to the first 5 pages, with 10 results on each page. The following websites were also searched with no restriction on page numbers of the search results: Canadian Institute for Health Information (CIHI);^31^ National Collaborating Center for Determinants of Health;^32^ and Ontario Public Health Libraries Association (OPHLA)-Custom Search Engine for Canadian Public Health information.^33^ For this study we also partnered with the Urban Public Health Network (UPHN), an association of urban local public health units in Canada; in order to ensure coverage of their membership, we also included their websites in our grey literature search (see Appendix B) with no restriction on the page numbers of the search results. The keywords searched to capture relevant grey literature included “avoidable mortality,” “avoidable death,” “preventable mortality,” “preventable death,” “amenable mortality,” and “amenable death.” We did not include the keyword “premature death” as we were focused on avoidable mortality.

### 2. Screening and identifying relevant studies

All the articles were imported into Rayyan^®^ for deduplication and title and abstract screening. The first author reviewed the articles by title and abstract to find relevant studies using the inclusion-exclusion criteria in Table 1. We were only interested in articles investigating socioeconomic status indicators as they are amongst those most frequently implicated as a contributor to inequities in health.^34^ In assessing socioeconomic status, researchers often consider factors such as income, education, employment, and occupation, analyzing them either individually or combining them into a single indicator. These indicators are usually measured at the household or area level so that they can relate to children’s health outcomes as well. We excluded articles investigating a specific cause of death or premature mortality since they are different from indicators of avoidable mortality. As we were interested in studies investigating the entire avoidable mortality at-risk population (aged 0 to 65 or 75), we excluded articles that limited their analysis to only a subset of this population, such as one sex, race, or children. The strict inclusion-exclusion criteria and minimal risk of misclassifying articles led us to use a single reviewer for screening and selecting the articles.

**Table 1.** Inclusion and exclusion criteria for selecting the articles and grey literature.

| Inclusion criteria |
| --- |
| (1) Articles which quantitatively evaluated socioeconomic inequalities in any type of avoidable mortality, defined as premature mortalities which could have been potentially avoided through primary, secondary, or tertiary prevention measures. |
| (2) Articles whose socioeconomic status of interest are one or more of income, education, and employment/occupation indicators. |
| Exclusion criteria |
| (1) Articles investigating a specific cause of death (e.g. cancer mortality). |
| (2) Articles investigating premature mortality without mentioning avoidable/preventable/amenable mortality. |
| (3) Articles that did not investigate the entire avoidable mortality at-risk population (i.e., 0 to 65 or 75 years of age,) assessing only a sub-sample of the population at-risk (e.g., recruitment of participants in a case-control study, a specific sex, children) |

### 3. Selecting eligible studies

After screening the titles and abstracts, the selected articles were transferred from Rayyan® to Paperpile® for in-depth, full-text review. At this stage, the first author evaluated the complete texts using the inclusion-exclusion criteria, determining the final article selection. Once all articles from the searched databases were assessed, the first author applied the criteria outlined in Table 1 to evaluate results from the grey literature search and decide on their inclusion in the review. Finally, the reference lists of all chosen articles were examined to identify any additional pertinent studies.

### 4. Charting the data

We created a spreadsheet to catalog characteristics and data from the chosen articles and grey literature. This form captured details such as the study’s timeframe, design, region examined, unit of analysis, population, SES indicators, definition of avoidable mortality, upper age boundaries, avoidable mortality measurement methods, and techniques used to analyze the relationship between avoidable mortality and SES. Additionally, we documented findings regarding the association between SES and avoidable mortality, the International Classification of Diseases (ICD) codes employed to identify avoidable deaths, and the classification of Ischaemic Heart Disease (IHD).

### 5. Collating, summarizing, and reporting the results

Finally, we compiled, summarized, and presented the gathered data. We highlighted the definitions of avoidable mortality used across the articles and determined whether Ischaemic Heart Disease was considered an avoidable cause of death. We summarized the insights from the articles regarding SES disparities in avoidable mortality and identified the SES indicators employed. Moreover, we compared the list of avoidable mortalities of a sample of five articles that used different definitions of avoidable mortality to identify avoidable deaths.

## Results

### Search results

In total, we identified 2,457 articles. Of these, 2,436 articles were obtained by searching the three predetermined databases using our tailored syntax (see Appendix A). From grey literature, we found 16 articles, and 5 more articles were identified by examining the reference lists of the included studies. The database search yielded 810 articles from Ovid Medline, 669 from Web of Science, and 957 from Scopus. After removing duplicates, 1,393 potential records underwent title and abstract screening. Of these, 1,301 were deemed irrelevant to our research and consequently excluded. This left 92 articles for a full-text review. From these, 58 were further excluded with the reasons for exclusion documented. Figure 1 offers a PRISMA flowchart detailing the selection process of this scoping review.

**Figure 1.**
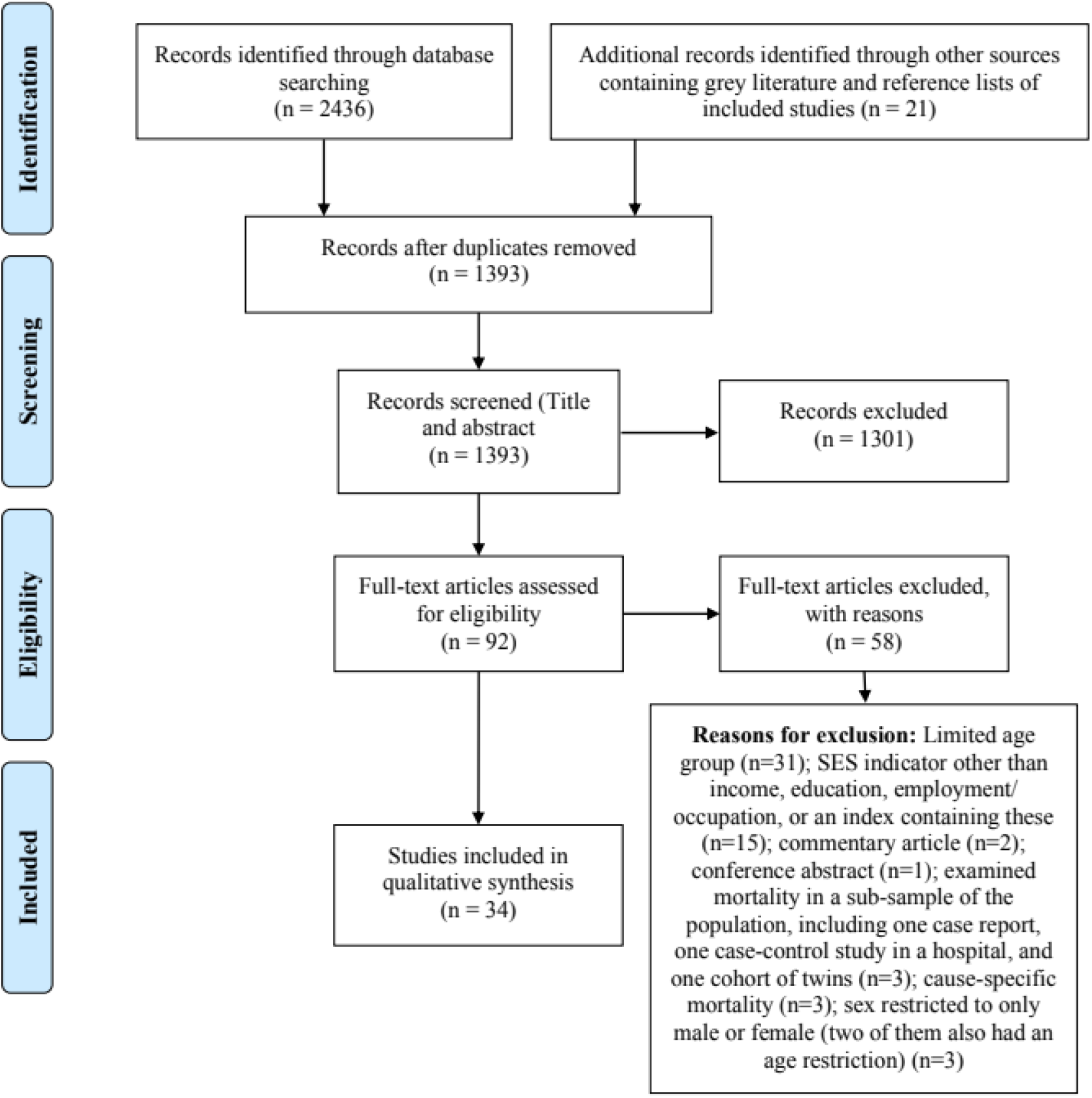
PRISMA flow diagram.

The grey literature search results were as follows: the World Health Organization (0 records); Public Health Agency of Canada (0 records); United Nations (1 record found and excluded after screening); OECD (1 record found and excluded after screening); CIHI (2 records found, 1 was included after screening); National Collaborating Center for Determinants of Health (1 record found and excluded after screening); OPHLA-Custom Search Engine for Canadian Public Health information (6 records found and 3 were included after screening). Additionally, we identified 5 records from the UPHN members’ websites, of which one report was considered suitable for inclusion after the screening process.

In conclusion, 34 articles were selected for detailed review and data extraction, comprising 29 scientific papers and 5 grey literature documents.

### Study designs and distribution by continent and timeline

The majority of the chosen studies originated from European nations, specifically England, Finland, Spain, France, Switzerland, and Hungary. The geographical distribution of the selected studies by continent is presented in Figure 2. Note that the grey literature is not considered in Figure 2 since our grey literature search was primarily centered on Canadian sources.

**Figure 2.**
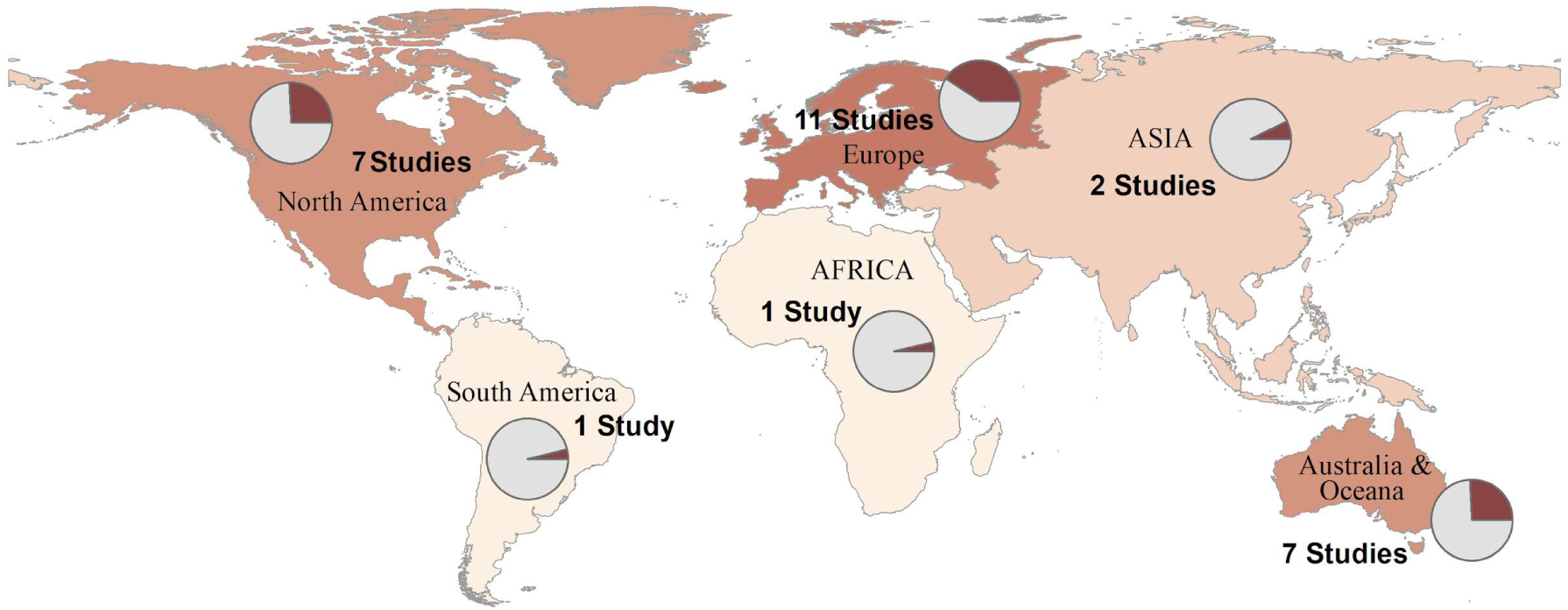
Geographical distribution of included studies (excluding grey literature).

Studies of avoidable mortality heavily rely on having access to previously collected death data. Analyzing and disseminating this data demands time, so the study coverage years might not always match their publication dates. Figure 3 contrasts these trends, showcasing the years studied in blue and publication years in orange. A noteworthy observation is that a significant portion (7 out of 34 articles) were unveiled in 2019, with 4 classified as grey literature. Moreover, there’s been a rising trend in the frequency of data collection annually from 1971 to 2016.

**Figure 3.**
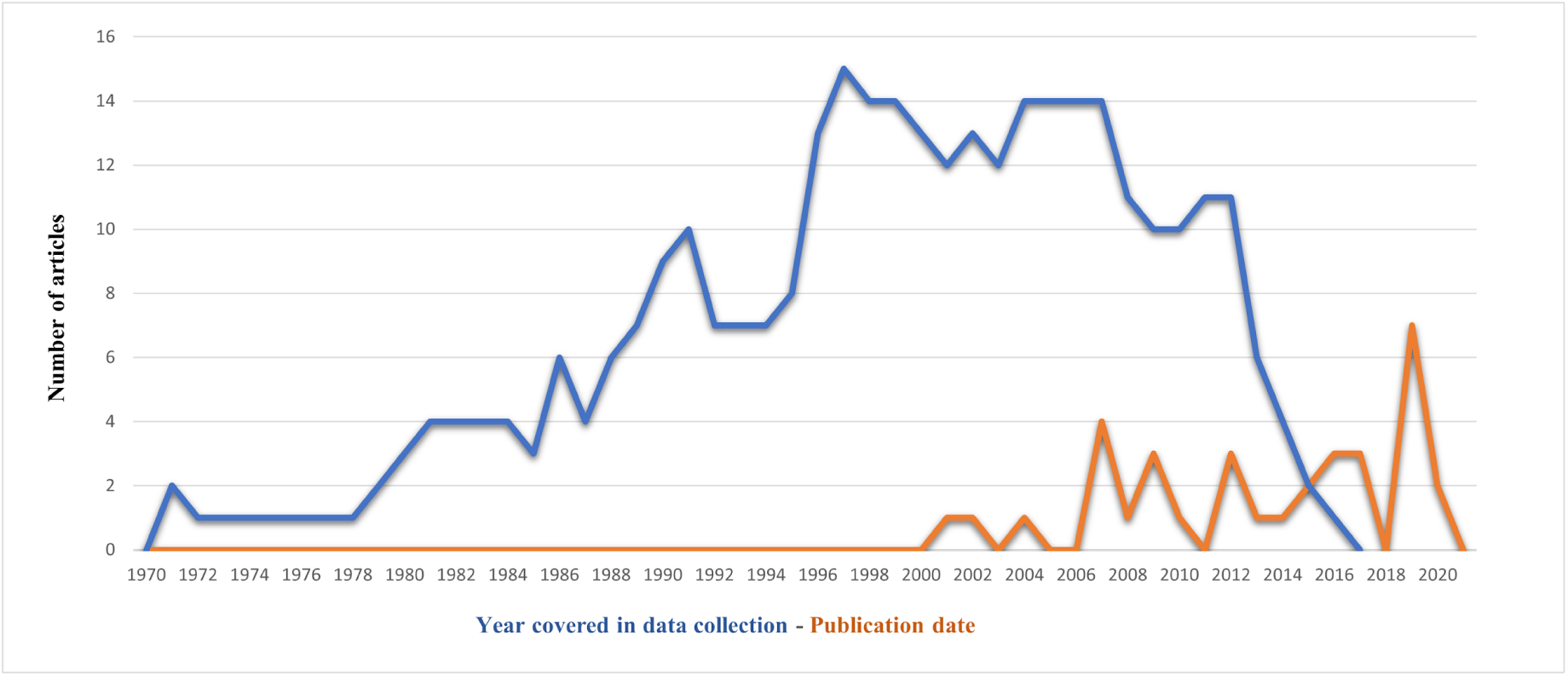
Annual data collection frequency in selected studies (blue line) and publication dates (orange line).

Most of the selected studies adopted quantitative observational approaches. To explore disparities in avoidable mortality, researchers employed a myriad of designs such as population-based, ecological, longitudinal, cross-sectional, prospective cohort surveys, exploratory spatial analysis, and official reports. The majority established an upper age threshold of 75 years. However, three distinct studies concentrated solely on deaths before age 65, covering the spans of 1971-2008, 1997-2001, and the 1990s, respectively.^35–37^

### Unit of analysis

The “unit of analysis” pertains to the level of grouping at which researchers measured avoidable mortality, subsequently stratifying these measurements based on SES to examine disparities. Most studies used small areas as their unit of analysis although the size of these areas varies between concepts and countries. These areas include the Lower Super Output Area (LSOA) in England,^38,39^ Dissemination Area (DA), ^8,17,39–42^ health region,^43^ and Census Tract (CT) in Canada^44^ and Spain,^19,20^ commune and canton in France,^36,37^ Local Government Area (LGA)^22,45^ and Statistical Local Area (SLA)^46,47^ in Australia, meshblock in New Zealand,^48^ districts in Brazil,^49^ neighborhoods,^21,50–52^ and other small areas.^11,18,53^

Only two studies used individual-level data for their analysis.^54,55^ A study carried out in Taiwan examined inequality between townships (city districts),^35^ and three articles investigated SES inequality in avoidable mortality at the municipal level.^56,57^ Surenjav et al. examined both provincial and municipal (capital) data,^58^ and Neethling et al. compared provinces and population groups in their analysis.^59^ Table 2 presents a summary of the selected articles’ characteristics.

**Table 2.** Summary of the selected articles.

| First author, year | Data years | Study design | Study region | Unit of analysis | SES<sup>1</sup> indicator | Findings (association between SES and AM<sup>2</sup>) |
| --- | --- | --- | --- | --- | --- | --- |
| ASIA |  |  |  |  |  |  |
| Chen, 2016 | 1971-2008 | longitudinal | Taiwan (354-358 townships) | township (city district) | mean annual household income | **Aggregated data:** In every period, the lowest income quartile had the highest age-standardized mortality rates, followed by areas in the second lowest, second highest and highest income quartiles. **Time trend:** Amenable mortality fell consistently from 1971-75 to 2006-08 across virtually all income quartiles, but fell faster in the more affluent townships than the less affluent townships. |
| Surenjav, 2016 | 2007-2014 | longitudinal | Mongolia | provinces /capital | percent of poor households | **Aggregated data:** Bivariate analysis did not reveal any significant association between crude AM and percent of poor households. |
| AFRICA |  |  |  |  |  |  |
| Neethling, 2019 | 1997-2012 | population-based retrospective | South Africa | province and population groups | Because of the existence of inequalities in education, income and welfare along racial lines due to the legacy of apartheid, apartheid classification was used as a proxy for SES. | **Aggregated data:** In 2012, there were huge inequalities in amenable mortality between different population groups. The white population had a much lower proportion of amenable mortality (26.3%) compared with other population groups (>48%), while the African population had the highest proportion (64.8%). **Time trend:** A disparity in age-standardized amenable mortality rate by population group in 2000 was evident, which widened each subsequent year until 2005 and narrowed thereafter. The African population had the highest age-standardized amenable mortality rate of all population groups over the period, which was 4.6 times that of the white population in 1997, increasing to 6.5 times in 2005. |
| EUROPE |  |  |  |  |  |  |
| Asaria, 2016 | 2004/05-2011/12 | whole-population longitudinal | England | LSOA<sup>3</sup> | IMD<sup>4</sup> 2010 | **Aggregated data:** There were substantial socioeconomic gradients in amenable mortality indicators in 2004/2005. **Time trend:** Although amenable mortality indicators were improved between 2004/2005 and 2011/2012, the socioeconomic gradient was persistent. |
| Cookson, 2017 | 2004/05-2011/12 | whole-population longitudinal | England and Ontario | LSOA (England), DA<sup>5</sup> (Ontario) | IMD 2010 (for England), ON-Marg<sup>6</sup> 2006 (for Ontario) | **Aggregated data:** A socioeconomic gradient was consistently observed for both jurisdictions in each year of the study period, with higher amenable mortality in more deprived areas. |
| Feller, 2017 | 1996-2010 | longitudinal | Switzerland and 16 high-income countries | neighbourhood, country | Swiss-SEP<sup>7</sup> | **Aggregated data:** There was substantial socioeconomic inequalities in amenable mortality. All types of mortality showed increasing hazards with lower socioeconomic position. |
| Hoffmann, 2014 | 1995-2009 | ecological | 15 large cities in Europe | small area | deprivation index | **Aggregated data:** All statistically significant rate ratios indicate a positive association between area deprivation and AM. |
| Manderbacka, 2013 | 1988-2007 | register-based cohort | Finland | individuals | household disposable income | **Aggregated data:** Mortality amenable to health policy measures contributed 35 per cent (4 years) to socio-economic differences in life expectancy at age 35. Mortality amenable to health care contributed 17 per cent (0.7 years) to the total difference between the highest and lowest income deciles. |
| Nagy, 2012 | 2004-2008 | ecological | Hungary | municipality | Hungarian specific deprivation index | **Aggregated data:** Strong positive association was found between the risk of mortality amenable to health care and deprivation index. |
| Nolasco, 2009 | 1996-2003 | transversal ecological | Alicante, Castellon, and Valencia (Spain) | CT<sup>8</sup> | deprivation index | **Aggregated data:** In all the three cities, the rate ratio of death for men in the least vs. highest privileged socioeconomic level was over 2; for women, these differences are only statistically significant in the city of Valencia. |
| Nolasco, 2015 | 1996-2001, 2002-2007 | ecological | 33 Spanish cities | CT | SES variable | **Aggregated data:** The estimated rate ratios show the excess risk of death at the lowest level of SES compared to the highest level. |
| Rey, 2009 | 1997-2001 | cross-sectional | France | commune<sup>9</sup> | deprivation index (FDep99)<sup>10</sup> | **Aggregated data:** Avoidable mortality was much more strongly associated with the FDep99 index (+77% between the fifth and the first quintile), than the other causes for the same age group (+32%). |
| Windenberger, 2012 | 1990s | longitudinal ecological | France | commune' and 'canton' | deprivation indexes (FDep) | **Time trend:** In 1988-92, the 'avoidable' mortality was 40% higher for the fifth deprivation quintile communes than for the first quintile communes (78% higher in 1997-2001). The increase in the association between the two periods was high for all the 'avoidable' causes of death considered separately. |
| Weisz, 2008 | 1988-90, 1998-2000 | cross-sectional | Paris, Inner London and Manhattan (France, England and Wales, US) | neighbourhood | pre tax average household income (for Paris and Manhattan), deprivation index (for London) | **Aggregated data:** The results from OLS<sup>11</sup> regression suggests a correlation between neighbourhood-level income and percentage of AM, during 1998-2000, in Manhattan at the 1% level, but no significant correlation in Paris or London at the 5% level. Despite this correlation, negative binomial regression results reveal that residence in a low income neighbourhood, as compared to the remainder of the city, is significantly correlated with increased AM rates per 1000 population in all three urban cores. |
| OCEANIA |  |  |  |  |  |  |
| Hayen, 2002 | 1980-2000 | longitudinal | New South Wales, Australia | LGA<sup>12</sup> | IRSD<sup>13</sup> | **Trends in time:** Rates of PAM<sup>14</sup> have decreased steeply for the three SES groups. However, the decrease has been more rapid for the highest SES group. Therefore, there was an increased relative gap between the highest SES group and the two lower SES groups. By contrast, the relative gap between the lowest and middle decreased slightly for males and remained almost constant for females. |
| Butler, 2010 | 2002-06 | exploratory spatial analysis | Australia | SLA<sup>15</sup> | a composite score of deprivation (remoteness areas, physician to population ratios, and IRSD) | **Aggregated data:** There was a strong relationship between the combined score of deprivation and AM. |
| Korda, 2007 | 1986,1991, 1997, 2002 | longitudinal | Australia | SLA | index of disadvantage (percentage of low-income families and percentage of early school leavers) | **Aggregated data:** The incidence rate differences and rate ratios comparing Q1 with Q5 in each year show significant absolute and relative socioeconomic inequality, respectively, in both avoidable and non-avoidable mortality. **Time trend:** The annual percentage decline in AM at the higher end of the socioeconomic continuum was larger than at the lower end, with increasing relative inequality between 1986 and 2002. The absolute inequality decreased between 1986 and 2002. |
| Piers, 2007 | 1979-2001 | longitudinal | Victoria, Australia | LGA | IRSD | **Aggregated data:** There was a clear gradient in total AM rates, highest in the most disadvantaged and lowest in the least disadvantaged IRSD quintile. |
| Tobias, 2001 | 1996, 1997 | cross-sectional | New Zealand | small area | NZDep96<sup>16</sup> index | **Aggregated data:** With few exceptions, all the conditions included in the AM category show a socioeconomic gradient in cause specific mortality. The gradient is steepest for Secondary Avoidable Mortality (SAM) and flattest for Tertiary Avoidable Mortality (TAM). |
| Tobias, 2007 | 2000-02 | cross-sectional | New Zealand | small area | NZDep2001 index | **Aggregated data:** Variations in amenable mortality make a substantial contribution to inequalities in mortality for socioeconomic (deprivation) groups. |
| Tobias, 2009 | 1981-84 to 2001-04 | population-based | New Zealand | meshblock (approximately 100 people) | income | **Aggregated data:** Absolute income inequality was found in all-cause mortality. Amenable mortality contributed 20.8% and 27.4% to income inequality in all-cause mortality in 2001-04, and 28.4% and 32.6% in 1981-04, for males and females respectively. |
| SOUTH AMERICA |  |  |  |  |  |  |
| Grafova, 2020 | 2003-2013 | ecological | São Paulo, Brazil | districts as proxies for neighbourhood | educational attainment and two measures of neighbourhood income | **Aggregated data:** In low-income districts, AM rate was more than 1.5 times greater than in high-income districts. Regarding Bolsa Familia and AM, no significant relationship was found. Increased literacy rates are predicted to improve significantly AM in high-income districts. **Time trend:** This difference is persistent during the 2003-2013, when AM rate declined across districts of different income levels. |
| NORTH AMERICA |  |  |  |  |  |  |
| Ronzio, 2004 | 1989, 1990, 1991 | cross-sectional ecological | central cities the United States | city | percentage in poverty | **Aggregated data:** Income inequality has a stronger correlation with mortalities attributable to preventable or immediate causes compared with all cause mortality. |
| Masters, 2015 | 1986-2006 | retrospective longitudinal | United States | individuals | educational attainment | **Aggregated data:** Larger education gradients in mortality risk were found for causes of death that are under greater human control than for less preventable causes of death. |
| James, 2007 | 1971, 1986, 1991, 1996 | longitudinal | Canada-metropolitan areas | neighbourhood (CT) | income | **Aggregated data:** In 1996, avoidable causes of death together accounted for about half (49.6%) of all income-related excess mortality among men, and 42% among women. |
| Khan, 2017 | 2002-2012 | population-based longitudinal | Ontario, Canada | DA | income and education | **Aggregated data:** There was a downward gradient in age-adjusted AM rates with increasing in the income and education level, for immigrants and long-term residents. |
| Young, 2019 | 2012-2014 | population-based | 18 northern regions, Canada | health region | education, employment, income | **Aggregated data:** There was a strong influence of SES on PAM, with lower PAM in higher levels of SES. The correlation for income was less strong than attained education and employment. |
| Zygmunt, 2019 | 1993-2014 | retrospective population-based | Ontario, Canada | neighbourhood (DA) | ON-Marg | **Time trend:** Avoidable mortality rates almost halved (48.6%) from 1993 to 2014. The inequity gap in AM rate ratio between the most and least marginalized quintiles widened for all marginalization dimensions. |
| Zygmunt, 2020 | 1993-2014 | retrospective population-based | Ontario, Canada | neighbourhood (DA) | ON-Marg | **Aggregated data:** Results from logistic regression shows decedents living in the most (Q5) materially deprived and residentially unstable neighbourhoods had significantly greater AM than those living in the least marginalized neighbourhoods (Q1). **Time trend:** Overall, AM rates were almost halved (48.6%) from 1993 to 2014. Compared with treatable AM, preventable AM contributed the greater proportion of all avoidable deaths, increasing from a ratio of approximately 1.5:1 in 1993 to approximately 2:1 in 2014. |
| GREY LITERATURE - CANADA |  |  |  |  |  |  |
| CIHI,<sup>17</sup> 2012 | 2005-2007 | annual report | Canada | neighbourhood (DA) | income | **Aggregated data:** For both preventable and treatable mortality, there were gradients in the rates by socio-economic group. Mortality rates were consistently higher among people living in the least affluent neighbourhoods, with rates gradually decreasing as socio-economic status increased. Socio-economic gradients were steeper for preventable mortality than for mortality from treatable causes. |
| Cui, 2019 | 2007-2016 | provincial report | Manitoba, Canada | NA | income | **Aggregated data:** There were strong relationships between income and potentially avoidable death rates in urban and rural areas in both time periods. In urban settings, the rate of potentially avoidable deaths for residents of the lowest income areas was about 3.7 times higher than residents of the highest income areas in T1 (2007-2011) and T2 (2012-2016). In rural settings, the rate of potentially avoidable deaths for residents living in the lowest income areas was about 2.2 times higher than for residents of the highest income areas in T2. |
| MLHU, 2019 | 2011-2012 | report | Middlesex-London, ON, Canada | neighbourhood | ON-Marg | **Aggregated data:** Significant differences in potentially avoidable mortality existed between populations living in neighbourhoods with differing levels of socio-economic status. **Time trend:** Inequalities between low socioeconomic status and high socioeconomic status persisted for all years between 2003 and 2012. |
| MLHU, 2019 | 2011-2015 | report | Middlesex-London, ON, Canada | neighbourhood | material deprivation | **Aggregated data:** Preventable mortality increased with each material deprivation quintile and varied significantly by material deprivation quintile on average from 2011 to 2015. |
| Rasali, 2019 | 2009-2013 | provincial report | British Columbia, Canada | DA | income, education, employment, social and material deprivation indices | **Aggregated data:** Analysis of rates by income, education, employment, social deprivation, and material deprivation showed declines in preventable premature mortality as socio-economic conditions improve. Rates for treatable premature mortality showed a similar pattern, except having smaller disparity ratios for the socio-economic dimensions. |

**Notes:** <sup>1</sup> Socioeconomic status, <sup>2</sup> avoidable mortality, <sup>3</sup> lower super output area, <sup>4</sup> index of multiple deprivation, <sup>5</sup> dissemination area, <sup>6</sup> Ontario marginalization material deprivation index, <sup>7</sup> area-based index of Swiss socioeconomic position, <sup>8</sup> census tract, <sup>9</sup> the smallest administrative unit in France, <sup>10</sup> French deprivation index, <sup>11</sup> ordinary least square, <sup>12</sup> local government area, <sup>13</sup> index of relative socioeconomic disadvantage, <sup>14</sup> primary avoidable mortality, <sup>15</sup> statistical local area, <sup>16</sup> New Zealand deprivation index, <sup>17</sup> Canadian institute for health information.

### Avoidable mortality definition

Researchers have used various definitions of avoidable mortality as the foundation for their studies. Table 3 displays the definitions adopted within the selected articles. While numerous studies adopted definitions proposed by other researchers or established organizations, several articles introduced their own distinct definitions of avoidable mortality, drawing from pertinent literature and consultations with expert panels.

**Table 3.**
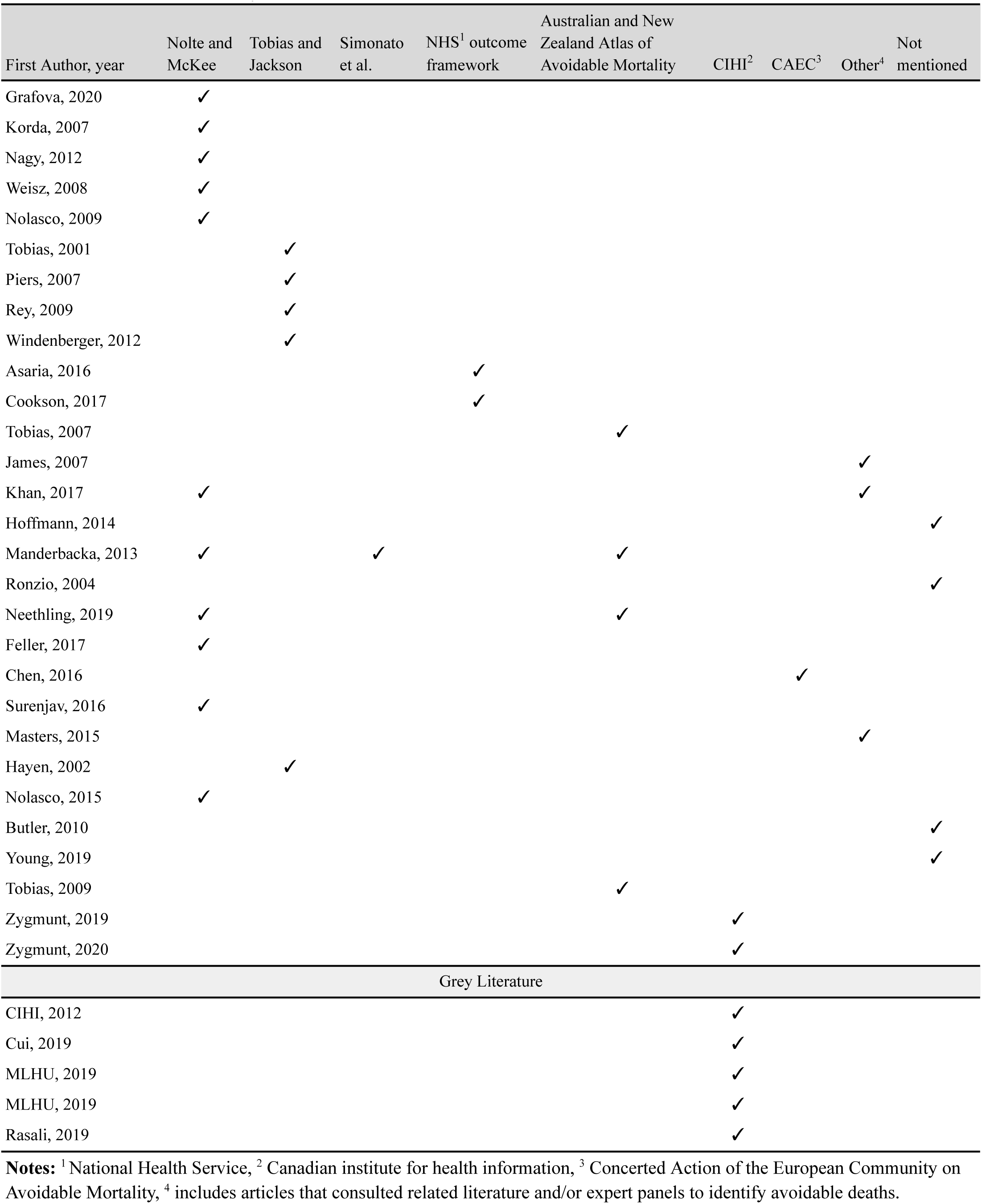
Avoidable mortality definition across the selected articles.

| First Author, year | Nolte and McKee | Tobias and Jackson | Simonato et al. | NHS<sup>1</sup> outcome framework | Australian and New Zealand Atlas of Avoidable Mortality | CIHI<sup>2</sup> | CAEC<sup>3</sup> | Other<sup>4</sup> | Not mentioned |
| --- | --- | --- | --- | --- | --- | --- | --- | --- | --- |
| Grafova, 2020 | ✓ |  |  |  |  |  |  |  |  |
| Korda, 2007 | ✓ |  |  |  |  |  |  |  |  |
| Nagy, 2012 | ✓ |  |  |  |  |  |  |  |  |
| Weisz, 2008 | ✓ |  |  |  |  |  |  |  |  |
| Nolasco, 2009 | ✓ |  |  |  |  |  |  |  |  |
| Tobias, 2001 |  | ✓ |  |  |  |  |  |  |  |
| Piers, 2007 |  | ✓ |  |  |  |  |  |  |  |
| Rey, 2009 |  | ✓ |  |  |  |  |  |  |  |
| Windenberger, 2012 |  | ✓ |  |  |  |  |  |  |  |
| Asaria, 2016 |  |  |  | ✓ |  |  |  |  |  |
| Cookson, 2017 |  |  |  | ✓ |  |  |  |  |  |
| Tobias, 2007 |  |  |  |  | ✓ |  |  |  |  |
| James, 2007 |  |  |  |  |  |  |  | ✓ |  |
| Khan, 2017 | ✓ |  |  |  |  |  |  | ✓ |  |
| Hoffmann, 2014 |  |  |  |  |  |  |  |  | ✓ |
| Manderbacka, 2013 | ✓ |  | ✓ |  | ✓ |  |  |  |  |
| Ronzio, 2004 |  |  |  |  |  |  |  |  | ✓ |
| Neethling, 2019 | ✓ |  |  |  | ✓ |  |  |  |  |
| Feller, 2017 | ✓ |  |  |  |  |  |  |  |  |
| Chen, 2016 |  |  |  |  |  |  | ✓ |  |  |
| Surenjav, 2016 | ✓ |  |  |  |  |  |  |  |  |
| Masters, 2015 |  |  |  |  |  |  |  | ✓ |  |
| Hayen, 2002 |  | ✓ |  |  |  |  |  |  |  |
| Nolasco, 2015 | ✓ |  |  |  |  |  |  |  |  |
| Butler, 2010 |  |  |  |  |  |  |  |  | ✓ |
| Young, 2019 |  |  |  |  |  |  |  |  | ✓ |
| Tobias, 2009 |  |  |  |  | ✓ |  |  |  |  |
| Zygmunt, 2019 |  |  |  |  |  | ✓ |  |  |  |
| Zygmunt, 2020 |  |  |  |  |  | ✓ |  |  |  |
| Grey Literature |  |  |  |  |  |  |  |  |  |
| CIHI, 2012 |  |  |  |  |  | ✓ |  |  |  |
| Cui, 2019 |  |  |  |  |  | ✓ |  |  |  |
| MLHU, 2019 |  |  |  |  |  | ✓ |  |  |  |
| MLHU, 2019 |  |  |  |  |  | ✓ |  |  |  |
| Rasali, 2019 |  |  |  |  |  | ✓ |  |  |  |

**Notes:** <sup>1</sup> National Health Service, <sup>2</sup> Canadian institute for health information, <sup>3</sup> Concerted Action of the European Community on Avoidable Mortality, <sup>4</sup> includes articles that consulted related literature and/or expert panels to identify avoidable deaths.

Avoidable mortality measures can be distinguished in two significant ways. First, there is a difference in how researchers phrase and conceptualize the type of mortality deemed avoidable. The term “amenable mortality” is mainly used to describe deaths that could have been averted through medical interventions. This term is often used interchangeably with “treatable mortality” in the literature.^12,60,61^ A definition by Nolte and Mckee, which is widely used in the selected articles (cited in 11 studies when identifying avoidable deaths,) characterizes “amenable mortality” as premature deaths that could have been avoided through timely and effective health care (i.e., secondary and tertiary prevention).^12^ Consequently, their list of amenable causes does not include deaths preventable via public health policy interventions^12^ which are referred to as “preventable mortality” and assessed by other researchers.^41,42,62^

Conversely, certain definitions, such as the one tailored for the Canadian setting, do not use the term “amenable mortality”.^8^ The Canadian Institute for Health Information (CIHI) categorizes “potentially avoidable mortality” into preventable and treatable mortality. Here, “preventable mortality” refers to deaths before the age of 75 that could have been potentially avoided through public health initiatives (i.e., primary prevention.) Meanwhile, “treatable mortality” pertains to premature deaths that could have been potentially avoided through health care interventions and treatments (i.e., secondary and tertiary prevention).^8^

Another important definition, not captured in our search, is that offered by the OECD.^63^ In 2022, the OECD and Eurostat partnered to create joint lists of “avoidable” causes of death.^63^ This definition builds upon the lists developed by Nolte and McKee,^12^ Eurostat,^64^ and CIHI,^8^ categorizing avoidable mortality into “preventable” and “treatable” mortalities. These categories are aligned with CIHI’s definitions.^63^ Within this framework, the term “amenable” from the prior Eurostat list has been rephrased as “treatable.” Similarly, the NHS Outcome Framework^65^ along with the Australian and New Zealand Atlas of Avoidable Mortality^61^ classify avoidable deaths into “amenable” and “preventable” categories, resonating with CIHI’s definition of “treatable” and “preventable” mortality.

On the other hand, Simonato et al.^13^ and Tobias and Jackson^11^ use unique terminology to define avoidable mortality. They partition avoidable mortality into three categories, each attributed to a set of mortalities that could be averted through one of the levels of prevention (i.e., primary, secondary, and tertiary levels). A detailed breakdown of the commonly used definitions in the selected articles is provided in Table 4.

**Table 4.** Description of frequently used definitions of avoidable mortality.

| Avoidable mortality Definition | Description | Subcategories of avoidable mortality |
| --- | --- | --- |
| Nolte and McKee | Amenable deaths are those that would not have occurred in the presence of effective health care (i.e., secondary prevention or medical treatment). | N/A |
| Tobias and Jackson | Avoidable deaths is partitioned among three subcategories. | (1) <u>Primary avoidable mortality (PAM)</u> constitutes conditions that are preventable, whether through individual behaviour change or population-level intervention (healthy public policy), i.e. primary prevention.<br>(2) <u>Secondary avoidable mortality (SAM)</u> constitutes conditions that respond to early detection and intervention, typically in a primary health care setting, as well as clinical preventive services such as screening, i.e. secondary prevention.<br>(3) <u>Tertiary avoidable mortality (TAM)</u> constitutes conditions whose case fatality rate can be significantly reduced by existing medical or surgical treatments, i.e. tertiary prevention. |
| Australian and New Zealand Atlas of Avoidable Mortality | This is an update on Tobias and Jackson's 2001 list. Avoidable causes of death represents those conditions whose associated mortality is substantially avoidable, given existing health and social systems in Australia, either through incidence reduction (prevention) or case fatality reduction (treatment) or a combination of both. Avoidable conditions are further classified into 'amenable' causes and 'preventable' causes. | (1) <u>Amenable causes</u> are defined as those causes whose case fatality could be substantively reduced by available health care technologies.<br>(2) <u>Preventable causes</u> are all other causes on the list, in that their associated mortality could be substantially reduced by preventing the condition from occurring in the first place, (i.e., incidence reduction). |
| Simonato et. al. | Avoidable causes are classified into three subcategories. | (1) <u>Causes avoidable through primary prevention</u> includes causes whose aetiology is in part attributable to lifestyle factors and/or to occupational risk factors. It also includes deaths from injury and poisoning, which are influenced in part by legal and societal measures such as traffic safety and crime reduction policies.<br>(2) <u>Causes amenable to secondary prevention through early detection and treatment</u> includes causes of death for which screening modalities have been established, as well as causes for which death is avoidable through early detection combined with adequate treatment.<br>(3) <u>Causes amenable to improved treatment and medical care</u> includes infectious diseases, deaths from which are 'avoidable' largely through antibiotic treatment and immunisation as well as causes that require medical and/or surgical intervention, deaths of which are related to complex interactions within the health care system, such as accurate diagnosis, transport to hospital, adequate medical and surgical care. |
| NHS outcome framework | This framework adopts the definition of amenable mortality proposed by Office of National Statistics (ONS); avoidable deaths are all those defined as preventable, amenable, or both, where | (1) <u>Amenable mortality</u> are deaths (subject to age limits if appropriate) that should not occur in the presence of timely and effective health care.<br>(2) <u>Preventable mortality</u> are deaths (subject to age limits if appropriate) that could be avoided by public health |
|  | each death is counted only once.<br>Avoidable causes of death are classified into two subcategories. | interventions in the broadest sense. |
| Canadian Institute for Health Information | (Potentially) avoidable mortality is defined as premature mortality, i.e. death occurred before the age of 75, that could have potentially been avoided in the presence of timely and effective health care services and public health policies, that is, through all levels of prevention. Avoidable causes of death are classified into two subcategories. | (1) <u>Preventable mortality</u> includes deaths that could have been averted through primary preventions. Preventable mortality informs efforts for incidence reduction.<br>(2) <u>Treatable mortality</u> includes deaths that could have potentially been prevented through secondary and tertiary preventions. Treatable mortality informs efforts for case-fatality reduction. |

Second, measures of avoidable mortality vary with respect to their operationalization using ICD codes (e.g., whether IHD is classified as avoidable). We evaluated the list of avoidable mortalities from five articles chosen at random and found significant variances. Table 5 presents these five articles, each tagged with a unique article code. These article codes serve as references when comparing their respective lists of avoidable mortality by ICD codes, as detailed in Appendix C. These differences largely arise from the adoption of varied definitions of avoidable mortality. Another contributing factor is the timeframe in which these lists were created; perceptions of avoidability evolve over time due to advancements in health care services and public health.

**Table 5.** Articles chosen for comparison of their lists of avoidable causes of death.

| Article code | Article title | Avoidable mortality list basis | Author(s) | Publication date |
| --- | --- | --- | --- | --- |
| 1 | Avoidable mortality in New Zealand, 1981-97 | Based on Tobias and Jackson | Tobias, M; Jackson, G | 2001 |
| 2 | Trends in amenable deaths based on township income quartiles in Taiwan, 1971-2008: did universal health insurance close the gap? | Based on CAEC classification of Avoidable Deaths | Chen, Brian K; Yang, Y Tony; Yang, Chun-Yuh | 2015 |
| 3 | Avoidable mortality by neighbourhood income in Canada: 25 years after the establishment of universal health insurance | The list of avoidable deaths was created with reference to classification lists from other studies (Charlton, etc) | James, Paul D; Wilkins, Russell; Detsky, Allan S; Tugwell, Peter; Manuel, Douglas G | 2007 |
| 4 | Trends and socioeconomic inequalities in amenable mortality in Switzerland with international comparisons | Based on Nolte and McKee | Feller, Anita; Schmidlin, Kurt; Clough-Gorr, Kerri M | 2017 |
| 5 | How much does health care contribute to health inequality in New Zealand? | Based on Australian and New Zealand Atlas of Avoidable Mortality | Tobias, M; Yeh, L C; Tobias, Martin; Yeh, Li-Chia | 2007 |

### Ischemic Heart Disease (IHD)

Despite the diverse views on which causes of death are deemed avoidable across the selected articles (see Appendix C,) Ischemic Heart Disease (IHD) remains a focal point in discussions about avoidability and its measurement. Among the chosen articles, there is no consensus on whether death due to IHD should be labeled as an avoidable cause. Numerous studies subdivide IHD into two or even three subsets of avoidable mortality. Given that both prevention and treatment are equally effective in avoiding IHD-related deaths, some studies — particularly those adhering to the CIHI definition of avoidable mortality^8^ — allocate half of the IHD-related deaths to treatable mortality and the remaining half to preventable mortality.^8,17,51,52,66^

In line with this, several studies included only half of the IHD mortality in their calculation of amenable mortality rates as proposed by Nolte and McKee.^12,21,48,50,53^ A study led by Tobias and Jackson introduced three categories for avoidable mortality: Primary Avoidable Mortality (PAM), Secondary Avoidable Mortality (SAM), and Tertiary Avoidable Mortality (TAM.) They divided IHD-related deaths into 50% PAM, 25% SAM, and 25% TAM,^11^ and a subsequent study following their categorization treated IHD deaths similarly.^45^

Conversely, a few studies treated IHD deaths like other avoidable causes, considering any death due to IHD avoidable mortality.^22,35,45,58,59^ Three studies reported IHD deaths by placing them in a separate group from other avoidable causes.^44,54,55^ Additionally, some articles opted out of classifying IHD as an avoidable cause, excluding it from their calculations of avoidable mortality rates.^18,20,37^ Figure 4 illustrates the number of articles that followed each approach regarding IHD-related deaths.

**Figure 4.**
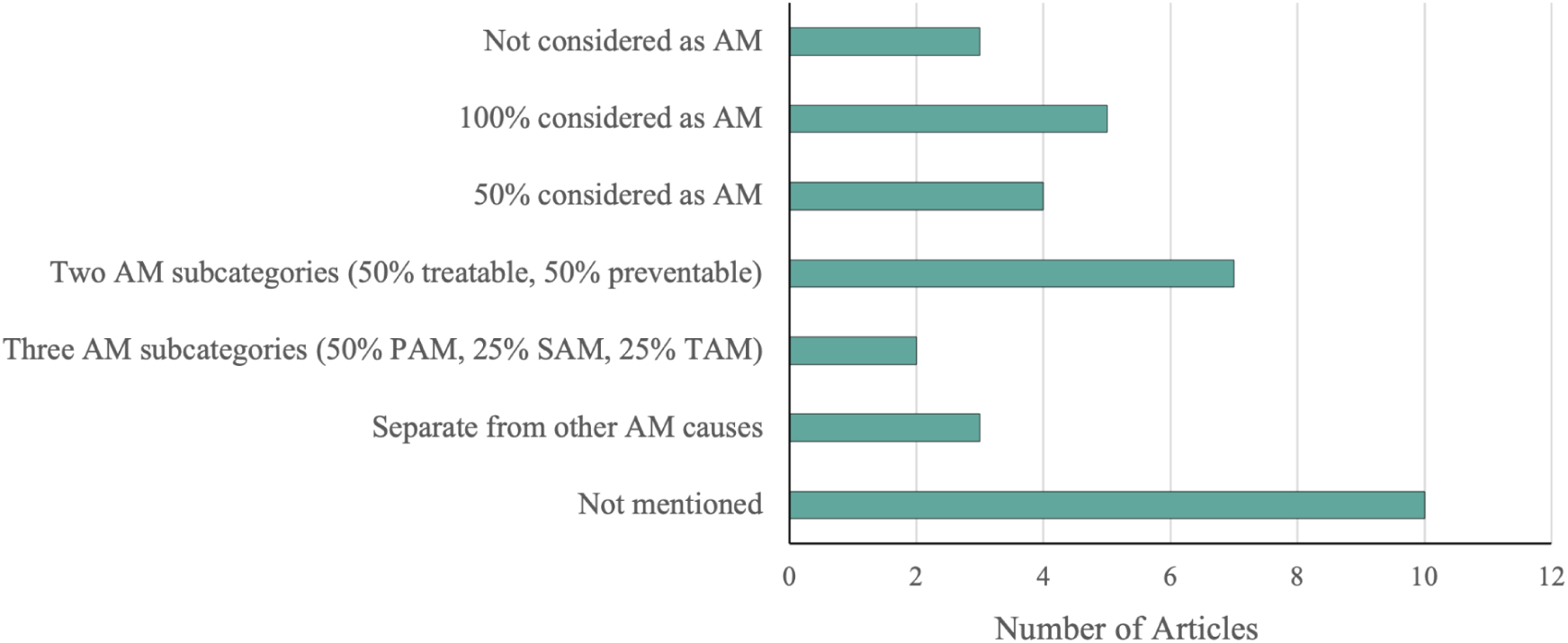
Categorization of Ischemic heart disease categorization as avoidable mortality.

### Socioeconomic status indicator

We classified the articles based on the type of indicator used for socioeconomic status (SES) into four categories, namely index, income, education, and employment. The index category includes any article using a predefined index for SES, such as the Ontario Marginalization Material Deprivation Index (ON-Marg),^39,51^ Index of Relative Socioeconomic Disadvantage (IRSD),^22,45,46^ and Index of Multiple Deprivation (IMD).^38,39^ Some authors constructed a variable representing SES in a population using indicators such as income and education.^19,20^ We placed these studies in the index group as well. To decide whether to include or exclude an article that used an SES index, we carefully examined the index to ensure that at least one of our indicators of interest (i.e., education, income, and employment/occupation) was taken into account in its construction. One study in South Africa used apartheid classifications (Africans, Whites, Asians, and Coloureds) as a proxy for socioeconomic status, arguing that income and education disparities continued to persist in the population even after apartheid was ended.^59^

Most of the articles (n = 20) used an index to investigate SES inequality in avoidable mortality. This number was 13 for income indicators, 4 for education, and 2 for employment. If a study examined more than one category in its analysis, we counted each category separately. For example, if an article examined income and education inequality in avoidable mortality separately, we counted this article once for the income category and once for the education category.

### Standardization for age and sex

Most of the included articles used directly age-standardized mortality rates to assess avoidable mortality. In some cases, mortality rates were also standardized by sex. However, some studies applied unique methodologies. For example, one study estimated smoothed standardized mortality rates using the Bayesian model proposed by Besag, York, and Mollie,^67^ to address the problem of age standardization in small areas.^18^ A Hungarian study calculated mortality amenable to health care ratios using full hierarchical Bayesian methods. In doing so, the authors calculated smoothed indirectly standardized mortality ratios using sex- and age-specific rates for the Hungarian population.^57^ A Canadian study calculated the age-standardized expected years of life lost (SEYLL) rate instead of the mortality rate, using the life expectancies of the richest income quintile as the standard.^44^ Another methodology examined the contribution of different groups of mortality, including death amenable to health care and death amenable to health policy, to life expectancy at age 35 and partial life expectancy between 35 and 75.^54^ Doing so, Manderbacka et al. were able to assess the impact of health policy and care on income disparities in life expectancy in Finland.^54^ In a study conducted by Masters et al., the authors tested the central claims of Fundamental Cause Theory (FCT) and then performed a retrospective cohort study to examine the association between preventable mortality and educational attainment.^55^ An ecological study conducted in France calculated the standardized mortality ratio by dividing the observed mortality in a spatial unit by the corresponding expected mortality.^36^

### Association between SES and avoidable mortality

The findings of the selected articles consistently indicate an association between SES and avoidable mortality rates, suggesting a decrease in avoidable mortality rates as socioeconomic conditions improve. This aligns with the substantial body of knowledge indicating the role of SES in population health.^2,3,23,68,69^ Khan et al. found a similar trend among immigrants and long-term residents, with a downward gradient in age-adjusted avoidable mortality rates as income quintiles increase.^40^ However, a study conducted in Mongolia reported no significant relationship between avoidable mortality rate and the percentage of poor households.^58^ This could be due to the lack of individual or neighborhood-level data in this study, which focused on the capital and provincial levels.^70^ Nevertheless, the study revealed higher amenable mortality rates in remote western provinces in Mongolia, characterized by harsh weather conditions, high poverty rates, lack of human resources for health, and poor infrastructure.^58^ Details on the findings of the selected articles are provided in Table 2.

While some studies adopted a descriptive approach and compared the avoidable mortality rates between different SES groups, many articles examined SES inequality in avoidable mortality using various types of analysis, including Poisson regression models, calculating Slope Index of Inequality (SII) and Relative Index of Inequality (RII), disparity rate ratio, incidence rate difference, random coefficient growth curve modeling approach, ecological regression, among other analytical methods.

All the studies that conducted a time trend analysis found a decline in avoidable mortality rates over time, while socioeconomic disparities persisted.^38,49,51^ A study conducted in South Africa found that the socioeconomic disparity widened from 2000 to 2005 and narrowed thereafter until 2012.^59^ Similar patterns were observed in France, with a 40% higher avoidable mortality rate for the fifth deprivation quintile communes compared to the first quintile in 1988-92, and 78% higher in 1997-2001.^37^ A study conducted in Taiwan reported a faster decline in avoidable mortality from 1971 to 2008 in affluent townships compared to less affluent townships.^35^ Similarly, an Australian study found a larger decline in avoidable mortality rates in higher socioeconomic groups, with increasing relative inequality and decreasing absolute inequality between 1986 and 2002.^47^

## Discussion and Conclusion

The findings of this review highlight the absence of a globally standardized terminology or definition when using the concept of avoidable mortality in health equity research. Common indicator definitions are essential for reliable comparison of health indicators within or between countries.^71^ While there’s a pressing need for standardized terminology and definition for avoidable mortality, it is important to recognize that a single, universal list of avoidable causes of death applicable in all countries and contexts is not likely feasible. The avoidability of specific causes of death may vary across countries with different levels of advancement in medical sciences. Therefore, it is crucial to consider the contextual factors and available healthcare technologies when developing lists of avoidable causes of death, acknowledging the variations across low-, middle-, and high-income countries. Nonetheless, this should not deter the establishment of a standardized definition, which is pivotal for supporting global comparability in this area of research.

The majority of studies captured in our scoping review were conducted in high-income countries, with Europe having the highest number of publications followed by Australia and New Zealand. This trend may be attributed to the presence of national or regional standard definitions for avoidable mortality in these countries, such as the well-established list of avoidable causes proposed for use in the Australian^61^ and European^72^ contexts. Nonetheless, it is worth noting that the availability of the list of avoidable causes of death primarily designed for high-income countries may hinder the conduction of avoidable mortality research in lower-income countries.

The impact of having an established list of avoidable causes of death on the number of conducted related studies can be observed not only in low-income countries but also in high-income countries like Canada. The introduction of Canada’s first list of potentially avoidable causes of death in 2012 by CIHI^8^ has led to a significant increase in inequality research on avoidable mortality in the Canadian literature.^8,17,51,52,66^ Four out of five Canadian governmental reports included in this review were published in 2019. These findings underscore the crucial role played by established definitions and lists of avoidable causes of death in promoting research on SES-related inequalities in avoidable mortality.

### Study limitations

A potential limitation of this scoping review is that only one reviewer screened the articles. However, given the rigorous inclusion-exclusion criteria we used, we are confident that the risk of misclassification of articles remains minimal. The scoping review also focused on studies of avoidable mortality which included analysis of SES-related inequalities. Studies which have not considered inequalities may have used additional conceptualizations and definitions of avoidable mortality.

## Data Availability

All data used in this study is public use and can be accessed through university libraries and the Data Liberation Initiative.

## Acknowledgments

This paper is based on a part of Anousheh Marouzi’s M.Sc. thesis and the first author thanks the advisory committee, Maureen Anderson and Sylvia Abonyi, for taking the time to assess her thesis and their insightful comments. The authors also thank Vicky Duncan and Thilina Bandara for reviewing earlier versions of this paper and for their helpful comments.

## Notes

**Conflict of Interest:** The authors declare that they have no conflict of interest.

### Competing Interest Statement

The authors have declared no competing interest.

### Funding Statement

This research was funded in part by the Urban Public Health Network. No other financial support was received for the research, authorship, and/or publication of this article.

