## Appendices for "Application of the Concept ‘Avoidable Mortality’ in Assessing the Socioeconomic Status related Inequalities in Health: A Scoping Review"

### Appendix A: Search Strategy.

| Database | Syntax |
|---|---|
| Medline | 1. (avoidable or preventable or treatable or amenable or untimely or unnecessary).tw.<br>2. death/ or mortality/ or "cause of death"/ or mortality, premature/<br>3. 1 and 2<br>4. (mortality or death* or mortalities).tw.<br>5. ((avoidable or preventable or treatable or amenable or untimely or unnecessary) adj (mortality or death* or mortalities)).tw.<br>6. 3 or 5<br>7. sociological factors/ or education/ or "social determinants of health"/ or exp socioeconomic factors/ or health care costs/ or health expenditures/<br>8. (socioeconomic* or socio-economic* or SES or "social determinant of health" or "social determinants of health" or education* or income* or salar* or wage* or employment or unemployment or job* or occupation* or poor or poverty or low-income or middle-income or "marital status").tw.<br>9. 7 or 8<br>10. 6 and 9<br>11. limit 10 to (english language and yr="2000 - 2020") |
| Web of Science | TS=((avoidable or preventable or treatable or amenable or untimely or unnecessary) NEAR/0 (mortality or death* or mortalities)) AND<br>TS=(socioeconomic* or socio-economic* or SES or "social determinant of health" or "social determinants of health" or education* or income* or salar* or wage* or employment or unemployment or job* or occupation* or poor or poverty or low-income or middle-income or "marital status")<br>AND LANGUAGE: (English)<br>Indexes=SCI-EXPANDED, SSCI, A&HCI, CPCI-S, CPCI-SSH, BKCI-S, BKCI-SSH, ESCI, CCR-EXPANDED, IC Timespan=2000-2020 |
| Scopus | (TITLE-ABS-KEY(avoidable or preventable or treatable or amenable or untimely or unnecessary) PRE/0 TITLE-ABS-KEY(mortality or death* or mortalities)) AND TITLE-ABS-KEY(socioeconomic* or socio-economic* or SES or {social determinant of health} or education* or income* or salar* or wage* or employment or unemployment or job* or occupation* or poor or poverty or low-income or middle-income or {marital status}) AND PUBYEAR > 1999 AND PUBYEAR < 2021 AND LANGUAGE(english) |

**Appendix B: Urban Public Health Network (UPHN) Members.**

| Health Authority | Region |
|---|---|
| Vancouver Island Health Authority | Victoria, B. C. |
| Vancouver Coastal Health Region | Vancouver, B. C. |
| Fraser Health | Surrey, B. C. |
| Alberta Health Services | Calgary + Edmonton, AB |
| Saskatchewan Health Authority | Saskatoon + Regina, SK |
| Winnipeg Regional Health Authority | Winnipeg, MB |
| Middlesex-London Health Unit | London, ON |
| Hamilton Public Health | Hamilton, ON |
| Ottawa Public Health | Ottawa, ON |
| Peel Public Health | Mississauga, ON |
| Region of York Public Health | York Region, ON |
| Toronto Public Health | Toronto, ON |
| Montreal Public Health | Montreal, QC |
| Monteregie Health and Social Services Agency | Longueuil, QC |
| Quebec City Health and Social Services Agency | Quebec City, QC |
| L'Agence de la sante et des services sociaux de laval | Laval, QC |
| Centre integre universitaire de sante et de services sociaux de l'Estrie | Estrie/ Sherbrooke, QC |
| New Brunswick- Saint John Area | Saint John, NB |
| New Brunswick-Fredericton Area | Fredericton, NB |
| Capital District Health Authority | Halifax, NS |
| Eastern Health | St. John's, NL |

**Appendix C: Comparison of Five Articles' Lists of Avoidable Causes of Death by ICD Codes.**

| Cause of Death | ICD-9 Codes | | | ICD-10 Codes | |
|---|---|---|---|---|---|
| Article code | 1 | 2 | 3 | 4 | 5 |
| Infections | | | | | |
| Selected invasive bacterial and protozoal infections | | | | | A38-A41, A46, A48.1, B50-B54, G00, G03, J02.0, J13-J15, J18, L03 |
| Intestinal infection | | | 001–009 | A00-A09 | |
| Tuberculosis | 010-018, 137 | 010-018, 137 | 010-018, 137 | A15-A19, B90 | A15-A19, B90 |
| Other (bacterial) infections (diphtheria, tetanus, poliomyelitis) | 023–031, 034–036, 084, 320, 3201–3209, 7700, 7711–2, 7714–9 | | 019–031, 034, 320–322, 381–383, 390–392, 680–686, 711 | A36, A35, A80 | |
| Poliomyelitis | | | 45 | | |
| Whooping cough | | | 33 | A37 | |
| Diphtheria | | | 32 | | |
| Tetanus | | | 37 | | |
| Measles | | | 55 | B05 | |
| Septicaemia | | | 38 | A40-A41 | |
| Ear infections | 381–383 | | | | |
| (Other acute) Respiratory infections | 460–466, 480, 487 | | 460–466 | | |
| Influenza | | | 487 | J10-J11 | |
| Pneumonia | | | 480-483, 485-486 | J12-J18 | |
| Immunisation-preventable | 032-033, 037, 045, 055-056, 3200, 7710,7713 | | | | |
| HIV/AIDS | 42 | | 42 | | |
| Sextually transmitted diseases | 090-099, 6140-6145, 6147-6169, 633 | | | | |
| Infections of the urinary system | | | 590-595 | | |
| Syphilis | | | 090-097 | | |
| Neoplasms | | | | | |

| | | | | | |
|---|---|---|---|---|---|
| Hepatitis and liver cancer | 070, 155 | | | | |
| Skin cancers | 140, 172, 173 | | 173 | C44 | |
| Melanoma of skin neoplasm | | | | | C43 |
| Non-melanotic skin neoplasm | | | | | C44 |
| Colorectal cancer | 153-154 | | | C18-C21 | C18-C21 |
| Oral cancers | 141, 143-6, 148-9, 161 | | | | |
| Lung cancer | 162 | 162 | 162 | | |
| Breast cancer | 174 | 174 | 174 | C50 | C50 |
| Cervical cancer (Malignant neoplasm of cervix uteri) | 180 | 180 | 180 | C53 | C53 |
| Stomach cancer | 151 | | | | |
| (Other) Uterine cancer (Malignant neoplasm of cervix uteri and body of the uterus) | 182, 179 | 182, 179 | 179, 182 | C54, C55 | C54, C55 |
| Cancer of bladder | | | | | C67 |
| Cancer of testis | 186 | | 186 | C62 | |
| Eye cancer | 190 | | | | |
| Thyroid cancer | 193 | | | | C73 |
| Hodgkin's disease | 201 | 201 | 201 | C81 | C81 |
| Leukaemia | 204 | | 204-208 | C91-C95 | |
| Lymphoid leukaemia- acute/chronic | | | | | C91.0, C91.1 |
| Benign cancers | 210–234 | | | | D10-D36 |
| Diseases of the Circulatory System | | | | | |
| Rheumatic fever/heart disease | 390-398 | | | | |
| Active rheumatic fever | | | 390-392 | | |
| Chronic rheumatic heart disease | | | 393-398 | I05-I09 | |
| Rheumatic and other valvular heart disease | | | | | I01-I09 |
| Hypertensive (heart) disease | 401–405, 4372 | | 401–405 | I10-I13, I15 | I11 |
| Hypertension and cerebrovascular disease | | 401–405, 430-438 | | | |
| Cerebrovascular disease | | | 430-438 | I60-I69 | I60-I69 |
| Ischaemic Heart Disease | 410-414 | 410-414, 429.2 | 410-414, 429.2 | I20-I25 | I20-I25 |
| Stroke | 431, 433, 434, 436 | | | | |
| Diseases of the Respiratory System | | | | | |

| | | | | | |
|---|---|---|---|---|---|
| Chronic obstructive pulmonary disease | 490-492, 496 | | 490-492, 496 | | |
| Asthma | 493 | 493 | 493 | | J45, J46 |
| All respiratory diseases (excl. pneumonia/influenza) | | | | J00-J09, J20-J99 | |
| Diseases of the Digestive System | | | | | |
| Peptic ulcer (Gastric and duodenal ulcer [ulcers]) | 531–534 | 531–534 | 531–534 | K25-K27 | K25-K28 |
| Acute abdomen | | | | | K35-K38, K40-K46 K80-K83, K85, K86, K91.5 |
| Appendicitis | 540–543 | 540–543 | 540–543 | K35-K38 | |
| Cholecystitis/lithiasis (and cholangitis) | | | 574-575.1, 576.1 | K80-K81 | |
| Pancreatitis | | | | | |
| Intestinal obstruction | 550–553, 560 | | | | |
| (Abdominal) Hernia | | 550–553 | 550–553 | K40-K46 | |
| Ileus without hernia | | | 560 | | |
| Gallbladder disease | 574, 57699 | 575-476 | | | |
| Cirrhosis of the liver | | | 571 | | |
| Diseases of the Genitourinary System | | | | | |
| Acute renal failure | 584 | | | | |
| Nephritis and nephrosis | | | 580-589 | N00-N07, N17-N19, N25-N27 | I12, I13, N00-N09, N17-N19 |
| Obstructive uropathy and prostatic hyperplasia | | | | | N13, N20, N21, N35, N40, N99.1 |
| Benign prostatic hyperplasia (Hyperplasia of the prostate) | | | 600 | N40 | |
| Infant and Maternal Causes | | | | | |
| Pregnancy complications (maternal mortality) | 630–632, 634–676 | 630–676 | 630–676 | O00-O99 | |
| Perinatal conditions (excluding stillbirths) | | 760-779 | 760-779 | P00-P96, A33, A34 | P03, P05-P95 |
| Low birthweight babies | 764-765, 769, 7707 | | | | |
| Sudden Infant Death Syndrome | 7980 | | | | |
| Neural tube defects | 740-742 | | | | |
| Newborn screening conditions | 243, 2552, 2701, 2711 | | | | |

| | | | | | |
|---|---|---|---|---|---|
| Congenital anomalies (Birth defects) | 743–7466, 7468–7479, 749–757 | | | | H31.1, P00, P04, Q00-Q99 |
| Congenital cardiovascular anomalies | | | 745-747 | Q20-Q28 | |
| Congenital digestive anomalies | | | 750-751 | | |
| Birth trauma and asphyxia | 767–768, 7701, 7720, 7723 | | | | |
| Other perinatal conditions | 766, 769, 7702–6, 7708–9, 7721–2, 7724–9, 773–779 | | | | |
| Unintentional Injuries | | | | | |
| (Motor vehicle accidents) Road traffic injury | 810-829 | | E810–825 | | |
| Accidents/Poisonings/Violence [Injuries] | | 800-999 | | | |
| Poisoning | 850-869 | | | | |
| Swimming pool injury | 8830, 9105, 9106 | | | | |
| Sport injury | 8840, 8845, 8860, 9170, 927 | | | | |
| Fire | 890-899 | | | | |
| Drowning | 910-9104,9107-9109,984 | | | | |
| Intentional Injuries | | | | | |
| Suicide | 950–959, 980–989 | | | | |
| Alcohol and Drug Use Disorders | | | | | |
| Alcohol related conditions | 291, 303, 3050, 4255, 5353, 5710-3 | | | | |
| Nutritional, Endocrine and Metabolic Disorders | | | | | |
| Nutrition | 260-9, 280, 281 | | | | |
| Deficiency anaemias | | | 280-281 | | |
| Thyroid disease | 240–242, 244 | | 240–246 | E00-E07 | E00-E07 |
| Diabetes | 250 | | 250 | E10-E14 | E10-E14 |
| Neurological Disorders | | | | | |

| | | | | | |
|---|---|---|---|---|---|
| Epilepsy | 345 | | 345 | G40, G41 | G40, G41 |
| Disorders of Musculoskeletal System | | | | | |
| Musculoskeletal infections | 680–686, 711, 730 | | | | |
| Osteomyelitis and periostitis | | | 730 | | |
| Adverse Effects of Medical and Surgical Care | | | | | |
| Iatrogenic conditions (Misadventure to patients during surgical and medical care) | 870–879 | | | Y60-Y69, Y83-Y84 | |

**Notes:** Grey cells indicate that the article did not consider the row-wise causes of death as avoidable.
